# Analyzing the Reversal Hypothesis of Obesity with Education: A Cross-Sectional Study of Adult Females in 34 Low- and Lower-Middle-Income Countries

**DOI:** 10.1101/19003004

**Authors:** Cynthia Y. Tang, Henok G. Woldu, Lincoln R. Sheets

**Affiliations:** School of Medicine, Institute for Data Science and Informatics, University of Missouri, Columbia, Missouri; Department of Health Management and Informatics, University of Missouri School of Medicine, Columbia, Missouri; Department of Health Management and Informatics, University of Missouri School of Medicine, Columbia, MO 65212, USA, United States of America

**Keywords:** Obesity, Social Determinants of Health, Education

## Abstract

**Setting:** Obesity, once considered an epidemic of the developed world, is now becoming an even more prominent problem than underweight in low and lower middle income countries (LLMICs). Ample literature has shown that as a country’s income increases, the burden of obesity shifts from the rich to the poor. This is known as the “Reversal Hypothesis.” Many studies have explored the effects of various social determinants of health on obesity, but few have studied education as an independent variable on female obesity across LLMICs.

**Objective:** Globally, adult females have a higher prevalence of obesity and the obesity shift occurs more quickly for women than for men. We aim to address this disparity and contribute towards the reversal hypothesis by exploring the association of education and obesity in women in LLMICs.

**Design:** In this cross-sectional study, we used a multi-national and multi-year database from the publicly available Demographic and Health Surveys program with data from 34 LLMICs. Education levels are standardized across countries during survey collection.

**Results:** Our age-adjusted prevalence ratio (AA-PR) analysis shows that women in LLMICs with higher education have a significantly greater prevalence of obesity than women with no education. We analyzed this phenomenon by individual nations, continents, and income classifications. Educated women living in low income countries are 5.12 times more obese than uneducated women (AA-PR, 95% CI=4.75, 5.53) and 3.42 times more obese in lower middle income countries (AA-PR, 95% CI=3.31, 3.54).

**Conclusion:** These findings highlight a need for more studies and policy attention focusing on female education levels, among other factors, to understand, predict, and prevent obesity in LLMICs.

**ARTICLE SUMMARY:** 

**Strengths and limitations of this study:**

- A rigorous sample size of 943,947 adult females in 34 LLMIC countries was utilized to study the association between adult female obesity and education level.
- Age-adjusted and age-and-wealth-adjusted prevalence ratios of obesity were analyzed based on 34 individual nations, three continents, and two major income categories.
- This study includes the most recent data available through the Demographic and Health Surveys program, which standardizes education levels during data collection, allowing for comparison between all surveyed countries.
- This study is limited by the relatively small number of countries for which data is available through the DHS dataset, and thus, further research will be needed to show whether these results are generalizable to other LLMICs.

## INTRODUCTION

The worldwide prevalence of obesity has nearly tripled since 1975.[1] Once an epidemic of developed countries, obesity is becoming increasingly problematic in low and lower middle income countries (LLMICs), surpassing the issue of underweight.[1;2] According to the Global Burden of Diseases, Injuries, and Risk Factors Study in 2017, high body mass index (BMI) has had a greater relative increase in exposure since 1990 than most of the other 83 risk factors that were evaluated.[3] Obesity is a primary risk factor for various medical conditions including ischemic heart disease and stroke, both of which are among the top 5 causes of deaths in LLMICs.[4-7] In 2015 alone, overweight and obesity were associated with 4.0 million deaths globally.[6] BMI is a modifiable risk factor, and thus, obesity is both a treatable and preventable condition. Understanding where and why it occurs is important in improving health outcomes.

It has been well studied that prior to 1989, obesity primarily affected those with high socioeconomic status (SES) in developing countries.[8] However, this trend has changed over the past few decades. In a 2004 review conducted on published data between 1989 and 2003, the burden of obesity in developing countries have begun to shift towards those with lower SES, particularly in women.[2] In 2019, Jaacks et al. proposed a conceptual model, termed the “Obesity Transition,” which highlights the shift of the burden of obesity from the rich to the poor as a country’s income level increases.[9] In low income countries, the poor are “protected” against obesity, while in high income countries, they are more susceptible. This phenomenon has been termed the “Reversal Hypothesis,” and the shift in obesity has been seen to be associated with a multitude of factors including but not limited to wealth, socioeconomic status, gender differences, ethnicity, urban and rural habitat, occupation, and education.[10-23] Furthermore, adult females have a higher prevalence of obesity, and the intranational shift of obesity from rich to poor occurs more quickly for women.[2;24] There is also country-level evidence that in countries transitioning from developing to developed status, income may be a risk factor, while education may be protective for obesity.[20;25;26] Thus, it is important to distinguish the individual effect of contributing factors, particularly between wealth and education on obesity.

There have been limited comprehensive studies focusing on an interpretable hypothesis testing models between obesity and education in the female population of LLMICs. Noteably, Kinge et al. explored educational inequalities from 70 countries of various gross domestic products (GDP) per capita from 2002-2013. In this study, we seek to continue building on the reversal hypothesis by focusing our exploration of obesity and education specifically in adult females across 34 LLMICs to determine the effect of education adjusted for age independently of other factors. We use the most recent data available through the Demographic and Health Surveys program ranging from 2008 to 2018 by analyzing the relationship by individual nations, global regions, and World Bank income classifications.

### Objective

This study aims to explore education as a factor in female obesity among 34 LLMICs. We hypothesize that education will follow the reversal hypothesis that unlike in high income countries, higher levels of education are associated with higher levels of obesity among adult women in LLMICs.

## METHODS

### Ethical Approval

Institutional ethical approval was not required to conduct this study. All data from the Demographic and Health Surveys (DHS) program is available to the public and anonymized.

### Population

Five inclusion criteria were used: (a) Demographic and Health Surveys (DHS) results were available since 2008, (b) the survey results included body mass index (BMI) values, (c) the survey results included education levels, (d) the country was classified by the World Bank as “low income” (Gross National Income per capita of less than 995 USD) or “lower middle income” (Gross National Income per capita between 996 and 3895 USD),[27] and (e) survey results were from women of reproductive age (ages 15 to 49). Women who were pregnant were excluded from analysis.

### Patient and Public Involvement

Patient and public involvement (PPI) were not directly included in this study. However, the DHS database used in the study was developed with PPI in the form of survey responses.

### Data Sources

Data were obtained from the Demographic and Health Surveys (DHS) program. The DHS program is a multinational and multi-year publicly available database funded by the United States Agency for International Development (USAID). The DHS program has collected data from more than 400 surveys in over 90 countries that are either classified as a developing country or countries that are receiving foreign aid from the United States. Sample weights were used by the DHS program to adjust for over- and under-sampling within different regions of each country to ensure proper representation. Surveys are generally conducted every five years. In addition, the highest level of education attended, but not necessarily completed, is reported. Educational categories are standardized between countries into the following categories: No education; primary, secondary, and high education.[28;29] Data from the most recent surveys available at the time of analysis were used for this study ranging from 2008 to 2018.

### Data Elements

The main outcome variable is body mass index (BMI). The World Health Organization (WHO) classifies obesity as BMI above 30 kg/m^2^.[30] Covariates including country name, urban or rural category, age, education level, and household wealth index were studied. Urban or rural category has been explored in our previous study, showing that urban disadvantage is associated with more obesity than rural disadvantage, and therefore is omitted in this study.[31] The income class of each country was based on the World Bank Atlas Method for the 2019 fiscal year.[32] Of the 836,721 surveys available, 109 contained missing records on BMI and education. Missing data was ommitted from analysis. Strengthening the Reporting of Observational Studies in Epidemiology (STROBE) reporting guidelines were used.[33]

### Statistical Methodology

In step one, we used descriptive statistics to explore the data for each of the the 34 countries. The mean (+/-SD) of BMI was calculated for each country along with the total number of women included in the analysis. BMI of greater than 60 kg/m^2^ were excluded from analysis as these values are most likely reported in error. In step two, the crude prevalence ratio of obesity by education level was caculated at the individual country, continent, and country income classification levels.

Finally, a generalized linear model with binomial distribution and logarithmic link function was used to estimate the age-and-wealth-adjusted prevalence ration (AWA-PR) and age-adjusted prevalence ratio (AA-PR) of obesity for females with higher than secondary, or high school equivalent, education and females with no education in LLMICs. Age was adjusted as a confounding factor due to its variability with different age groups.[34]

The 95% confidence interval (CI) of each prevalence ratio was presented. A two-sided 0.05 alpha level of significance was used to determine the statistical significance of the results. A p-value less than 1% was considered statistically significant. The data analysis for this paper was generated using SAS software Version 9.4 of the SAS System for Unix. Copyright © 2013 SAS Institute Inc. SAS and all other SAS Institute Inc. product or service names are registered trademarks or trademarks of SAS Institute Inc., Cary, NC, USA.

## RESULTS

A total of 34 countries fit our inclusion criteria. To analyze the relationship between education level and obesity in women in LLMICs, we began by searching the DHS database and found data covering 19 low income countries and 15 lower middle countries (Table 1). Using this dataset, we compared obesity in women with post-secondary level education to those with no education to calculate the age-and-wealth-adjust prevalence ratio (AWA-PR) and age-adjusted prevalence ratio (AA-PR) (Figure 1) for each country, with confidence intervals (CI). Household wealth index and education were found to be colinear variables. In this study, we are interested in determining the effect of education as an individual factor. Therefore, we calculated the prevalence ratio of obesity by education level after adjusting for both age and wealth, and after adjusting for age alone. Our results show that women in LLMICs with higher than secondary education had a greater prevalence of obesity than women with no education (AA-PR=3.55, CI=3.44, 3.67) (Table 2). The AA-PR was statistically significant (P-value < 0.01) in 29 of the 34 countries analyzed, with the exceptions being Bolivia, Comoros, Côte d’Ivoire, São Tomé and Príncipe, and Tajikistan. The highest disparity was seen in Burundi (AA-PR=27.36, CI=13.60, 55.06), and the lowest significant disparities are seen in Honduras (AA-PR=1.75, CI=1.51, 2.04) and Pakistan (AA-PR=1.50, CI=1.29, 1.75). This prevalence ratio remained statistically significant when we analyzed the cumulative populations of all three continents, both income classes, and all 34 countries. After adjusting for age, we found that highly educated women living in low income countries are 5.12 times more likely to be obese than uneducated women (CI=4.73, 5.53) and those living in lower middle income countries are 3.42 times more likely to be obese (CI=3.37, 3.60). Of the African nations, the average AA-PR was 4.98 (CI=4.66, 5.31); in Asia, the average AA-PR was 3.57 (CI=3.44, 3.72); and in Latin America, the average AA-PR was 1.69 (CI=1.51, 1.88). This prevelence ratio could not be calculated for the Kyrgyz Republic because the prevalence of obesity in women with no education was zero; for this reason, the Kyrgyz Republic was also excluded from the cumulative prevalence ratios for Asia, low-middle-income countries, and all countries.

**Table 1.**
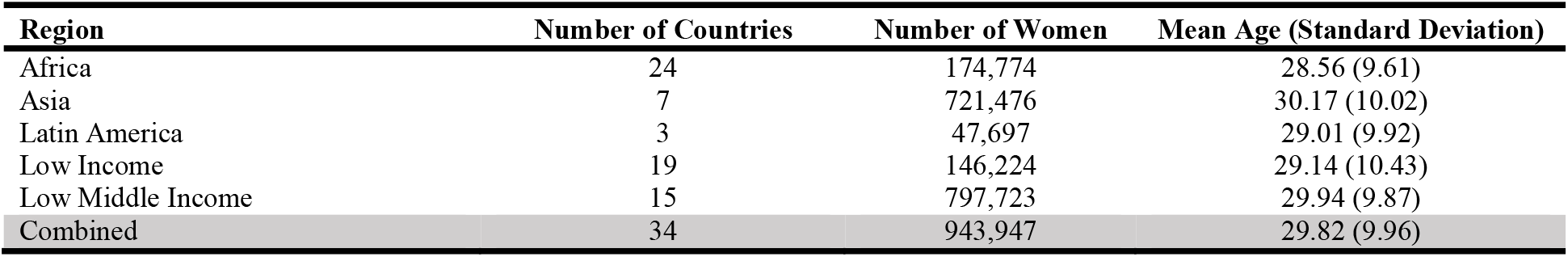
Demographics of 34 Low- and Lower-Middle-Income Countries.

**Table 2.**
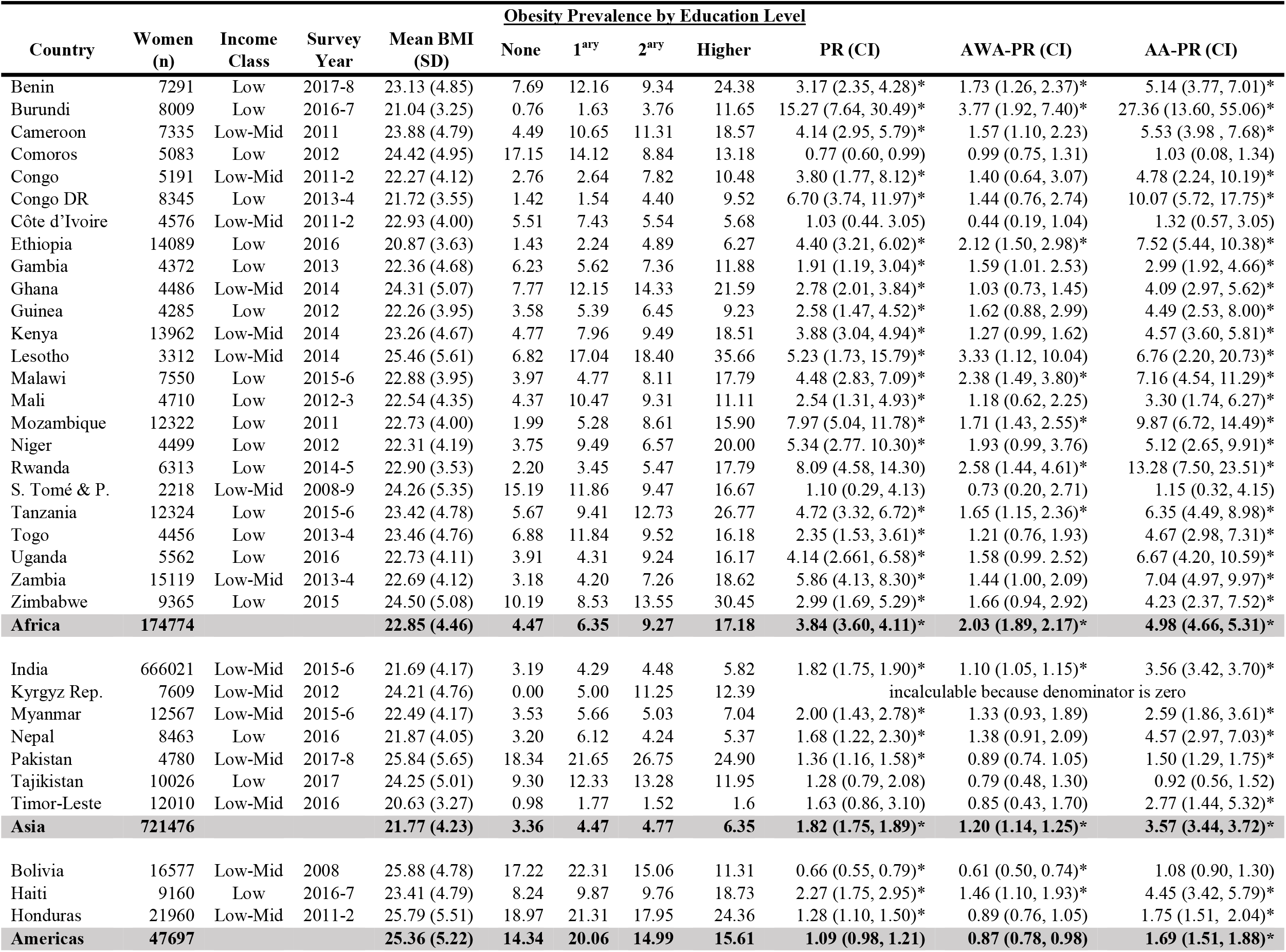
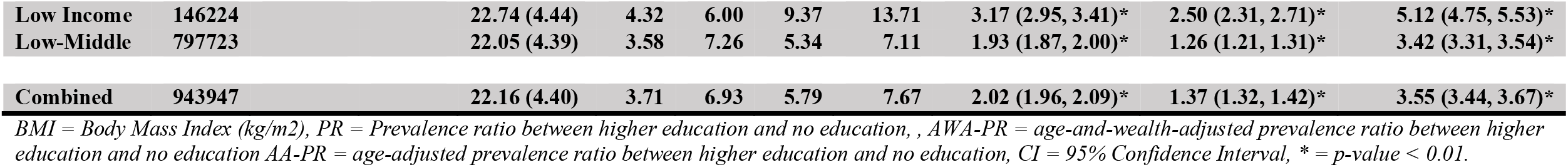
Age-Adjusted Obesity Prevalence By Education Level.

**Figure 1.**
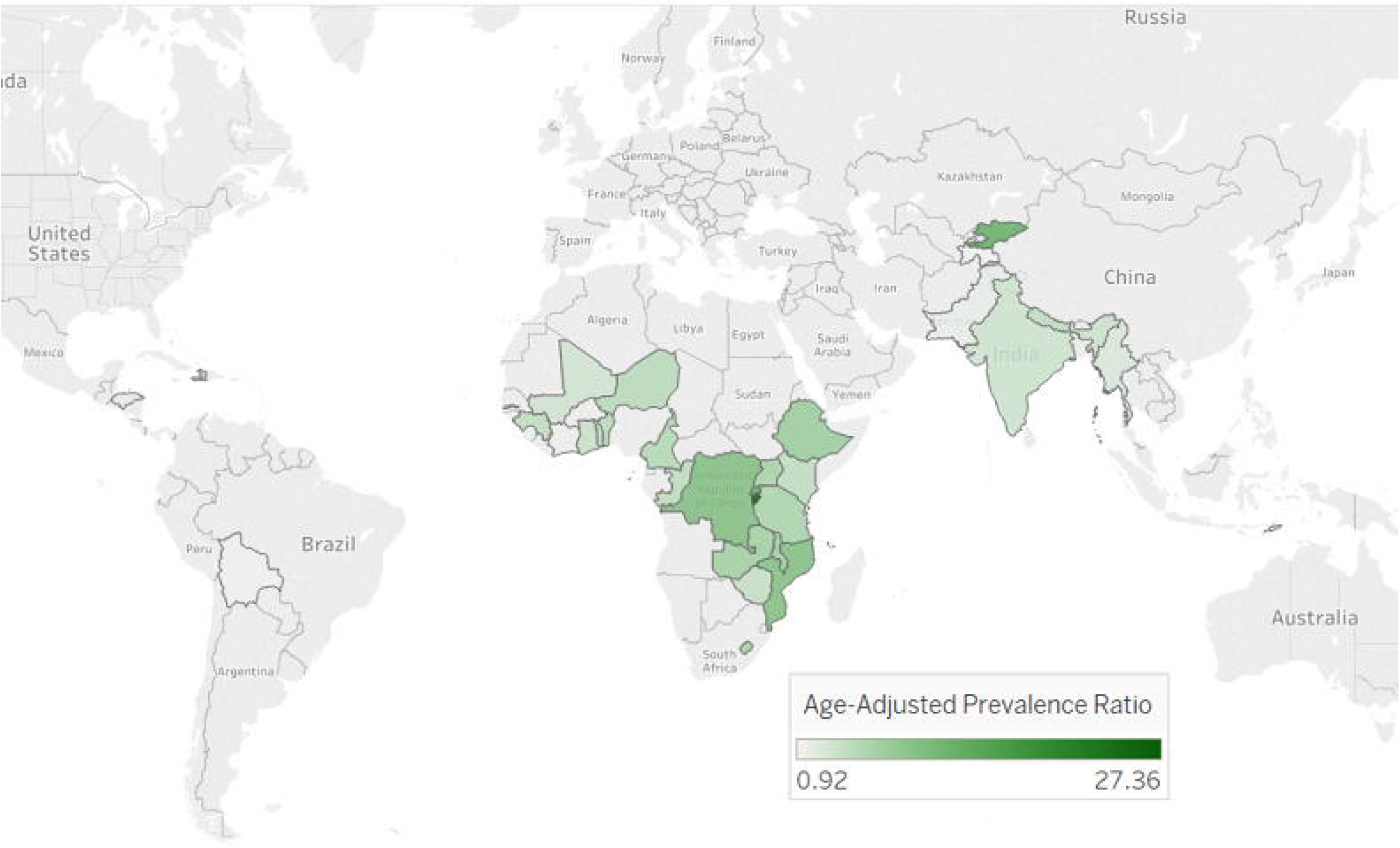
Geographical Representations of Age-Adjusted Prevalence Ratios of Highly Educated vs. Non-Educated Obesity by Country.

## DISCUSSION

The findings of this study support the reversal hypothesis that, contrary to the findings in high income countries, higher levels of education are associated with significantly higher levels of obesity among women in the majority of the analyzed LLMICs. We found that this association remained consistent even when adjusted for the major covariate of wealth. These findings are consistent with available literature.[11;16;20;35;36] The greatest disparities were found in Africa and in low income countries. Among the strengths of this study include the large sample size of nearly 1 million adult females in 34 LLMICs that were analyzed to explore the association between adult female obesity and education level. The data collection methods through the DHS are standardized across surveyed countries, allowing for reduced bias in the classifications and comparisons of education and other variables among different countries. Additionally, the study uniquely categorizes the analyses of age-adjusted prevalence ratios of obesity for each of the 34 countries from three continents, and two income classes.

The study is limited by the relatively small number of countries for which data is available through the DHS dataset, and thus further research will be needed to show whether these results are generalizable to other LLMICs. Additionally, our results cannot be generalized to higher income countries or to men in LLMICs, as we are complementing the existing literature studying similar relationships among those populations. The limited available data also does not allow us to account for intranational variability. It is important to note that BMI does not distinguish between muscle mass, bone density, and excess fat, and may vary based on body type and race. However, it is the current best standard for determining obesity and is the metric used by the WHO.[30]

Many factors may be contributing to the relationship found in this study. Higher levels of education may provide higher income opportunities, allowing for greater access to surplus food. Food scarcity is higher in LLMICs, possibly resulting in lower food intake in the less educated and presumably less wealthy population, thus “protecting” this population from obesity. Higher education may also provide job opportunities that are less manual labor-intensive and therefore, less energy expenditure is required. Because of this potential relationship between education and income potential, we identified household wealth index and education as colinear variables. The relative contributions of each factor could not be disentangled by our analysis. Further study is needed to determine the contribution of household wealth on obesity. Additionally, some countries have a cultural association between higher status with larger female body size, which may contribute to obesity in the more educated population as studied in countries including Ghana, Tanzania, and Uganda.[21;37;38] Studies have also shown discrepencies between self-perception of normal body weight versus actual overweight and obesity in developing countries.[39;40]

Finally, we acknowledge that there are many other factors contributing to the reversal hypothesis of obesity. While this study focused on education, factors including gender differences, ethnicity, urban and rural habitat, occupation, and salary are important to consider. These factors have been explored previously and leave room for additional opportunities for investigating the relative effect of these factors on female obesity in LLMICs in the simple hypothesis testing models and the more advanced predictive models. Even with these limitations, the evidence that obesity is more prevalent in women with higher education in LLMICs is an interesting and important step towards guiding future research in addressing and preventing female obesity.

High prevalence of obesity leads to adverse implications for obesity-related disabilities and mortality, societal health consequences, and economic consequences. Our results indicate that overall, more educated women in LLMICs face a higher prevalence of obesity. This knowledge may allow for political implementation of changes to counter this increase such as incorporating nutrition, accurate body-weight perceptions, and physical fitness into the education system. Clinicians can also play an invaluable role in educating female patients on proper nutrition, realistic body-weight perceptions, and increasing energy expenditure.

## CONCLUSIONS

The finding that obesity is significantly more prevalent in women with higher education is contrary to what is seen in high income countries. This provides strong support for the reversal hypothesis of obesity and highlights a need for more studies and public policy attention focusing on the reasons leading to the shift of the obesity burden in LLMICs. The multi-factorial relationship between female obesity and education in LLMICs requires additional investigation to determine the primary causes of this phenomenon and initiatives to address this problem. As obesity rapidly grows as a public health threat, understanding associations between obesity and education, particularly in women, will be invaluable in predicting and preventing the disorder in LLMICs.

## Data Availability

Data is publicly available through the Demographics and Health Surveys (DHS) Program. The DHS Program includes survey data on population, health, HIV, and nutrition. The database includes over 400 surveys spanning over 90 countries.

https://www.dhsprogram.com/

## ACKNOWLEDGEMENTS

The authors wish to thank the USAID for making their DHS data available to the public, Rachel Alexander for her help with formatting, and Genevieve Sheets for her editorial input on the manuscript.

## AUTHOR CONTRIBUTIONS

Cynthia Y. Tang, Henok G. Woldu, and Lincoln R. Sheets were involved in the study concept and design, acquisition, analysis, and interpretation of the data, and drafting and revision of the article.

## DATA AVAILABILITY

Data is publicly available through the Demographic and Health Surveys (DHS) Program. The DHS Program includes survey data on population, health, HIV status, and nutrition. The database includes over 400 surveys spanning over 90 countries. Data can be accessed at: https://www.dhsprogram.com/.

